# Subjective and Objective Assessment of Physical Activity in Older Adults: Results from Berlin Aging Study II

**DOI:** 10.1101/2022.11.17.22282445

**Authors:** Valentin Max Vetter, Duygu Deniz Özince, Jörn Kieselev, Sandra Düzel, Ilja Demuth

**Affiliations:** Charité – Universitätsmedizin Berlin, corporate member of Freie Universität Berlin and Humboldt-Universität zu Berlin, Department of Endocrinology and Metabolic Diseases (including Division of Lipid Metabolism), Biology of Aging working group, Augustenburger Platz 1, 13353 Berlin, Germany; Max-Planck Institut für Bildungsforschung; Charité – Universitätsmedizin Berlin, corporate member of Freie Universität Berlin and Humboldt-Universität zu Berlin, Department of Anesthesiology and Operative Intensive Care Medicine (CVK/CCM), Chariteplatz 1, 10117 Berlin, Germany; Charité – Universitätsmedizin Berlin(CBF), Department of Cardiology, Berlin, Germany; Berlin Institute of Health at Charité – Universitätsmedizin Berlin, BCRT - Berlin Institute of Health Center for Regenerative Therapies, Berlin, Germany

## Abstract

Physical activity has a substantial impact on health and mortality. Besides questionnaires that rely on subjective assessment of activity levels, accelerometers can help to objectify an individual’s physical activity.

Physical activity was assessed with the GT3X activity monitor (ActiGraph LLC, USA) in 797 participants of the Berlin Aging Study II (BASE-II) and compared with questionnaire data obtained from the Rapid Assessment of Physical Activity (RAPA).

Of all participants, 760 fulfilled the wear-time based inclusion criteria. Mean age was 75.6 years (SD: 3.8 years, range 66.0 – 94.1 years) and 53% of the included participants were female. Average wear time was 23.2 hours/day (SD 1.3 hours/day). Statistically significant differences between RAPA groups were found for all accelerometric variables except energy expenditure. Post-hoc analysis, however, suggested low agreement between subjective and objective assessment of physical activity. When focusing on the first question of the RAPA questionnaire, participants who reported to be “seldom or never active” showed statistically significant lower activity levels in Vector Magnitude, step counts, and cut-off defined moderate physical activity (p<0.05). Total cholesterol, high density lipoprotein cholesterol, low density lipoprotein cholesterol, triglycerides, fasting glucose and hemoglobin A1c were weakly correlated with accelerometric variables (Pearson’s r ≤ 0.25).

Results from this study suggest that subjectively assessed data from the RAPA and objectively assessed data from accelerometric variables obtained from the GT3X monitor are partially inconsistent in terms of the physical activity level of the participants.

## Introduction

The impact of physical activity (PA) on health and mortality is undisputed [1, 2]. Inactivity is estimated to be responsible for 9% of premature deaths in 2008 and between 6% and 10% of non-communicable diseases like type 2 diabetes, coronary heart disease and several forms of cancer [2]. The extent and prevalence of inactivity varies around the world, but on average, one-third of all adults do not achieve the PA levels recommended in public health guidelines [3]. To account for and examine activity related effects, a reliable assessment of overall PA in the past and on a daily basis is pivotal in epidemiological health studies. However, PA is a multidimensional process [4] and to date no gold standard for its assessment is available [5]. Questionnaires on PA impress with their simplicity, low costs, and easy repeatability but were shown to have limited validity when compared to other methods [5] and differ in the degree of resolution and time-frame of assessing PA. Another way of measuring PA is through accelerometers that assess more objectively the acceleration of the body part they are attached to (often in three axis). Decreasing costs, ease of use, and low invasiveness have made them a widely used tool for assessing PA even in large cohorts [6]. However, accelerometers produce an immense volume of data and different approaches to reduce it to one single metric (which would be needed for easy use in most subsequent analyses) are available [7]. Still, due to the complexity and multidimensionality of PA [4] there is no clear consensus which of these metrics would be best to measure overall activity [7-9]. Beyond that, it was shown that measurements differ in dependence on the body position where the monitor was worn [9-12], employed filters [13], data processing procedures and used algorithms for the aggregation of raw data (reviewed in [8]).

In this study, we analyze five PA measures obtained through the GT3X monitor and the ActiLife software package (including step counts, vector magnitude counts, and moderate PA) in 760 participants of the Berlin Aging Study II (BASE-II), which were assessed as part of the GendAge study [14]. Accelerometric variables were compared with self-rated PA, assessed through the Rapid Assessment of Physical Activity (RAPA), and correlations with age, BMI and blood parameters that are known to be associated with PA (e.g. total cholesterol (TC), high density lipoprotein cholesterol (HDL-C), low density lipoprotein cholesterol (LDL-C), triglycerides (TG), fasting glucose and hemoglobin A1c (HbA1c)) were examined.

## Methods

### Participants

A total of 1,100 participants originating from the Berlin Aging Study II (BASE-II) [15] aged ≥60 years (52.1% women) were re-invited and medically assessed (follow-up) as part of the GendAge study. The time interval between baseline examination and follow-up was on average 7.4 years [14]. All participants gave written informed consent. All medical assessments were conducted in accordance with the Declaration of Helsinki and approved by the Ethics Committee of the Charité – Universita□tsmedizin Berlin (approval number EA2/144/16) and were registered in the German Clinical Trials Registry as DRKS00016157.

### Actigraphy Raw Data Assessment and Data Processing with the Actilife Software Package

BASE-II participants were asked to wear the GT3X activity monitor (ActiGraph LLC, USA) like a wristwatch (i.e. usually their non-dominant wrist) for the (on average six) days that lay between the second and the third day of examination at follow-up. To increase the participants motivation to wear the device as long as possible they retrieved a summary report after they returned the accelerometers to the study site and were provided with instruction on how to wear the accelerometer correctly.

The devices were initiated before they were assigned to each participant according to the manufacturer’s instructions (default settings).

The manufacturers software package ActiLife 6 (ActiGraph LLC, USA) was used for data processing. Raw data was downloaded an aggregated into 10 second epochs. The low-frequency extension filter that increases the devices sensitivity to low-force accelerations was not applied to the data in this study [13]. Following the recommendations of Keadle and colleagues [16] and Choi and colleagues [17] the wear time validation algorithm by Choi implemented in the ActiLife software package was used to determine wear time of all participants. The algorithm derived wear time information was supplemented by the built-in wear time sensor of the GT3X-monitor. Arguello and colleagues [18] showed that the wear sensor has a lower sensitivity but a higher specificity in wear time detection than the implemented wear time validation algorithms. Therefore, conflicts between the Choi algorithm and the wear time sensor were manually solved following predefined rules:

- If the wear sensor indicates non-wear time and algorithm indicates wear time, non-wear time was chosen.
- If wear sensor indicates wear-time and algorithm indicates non-wear time, non-wear time was chosen.

If the results by wear time sensor or wear time algorithm were in obvious discrepancy with the measured accelerometric data, a decision by the evaluator (“wear time” vs “no wear time”) was made based on plausibility after screening the raw data. Periods of conflict shorter than 30 minutes were ignored, as their influence on the overall wear time estimation would be limited.

Biometric information such as age, sex, weight, and height were entered manually for each participant. Energy expenditure was calculated based on the algorithm by Freedson and colleagues [19] (“Freedson VM3 (2011)” option in ActiLife 6) and cut points for intensity of PA were defined by Keadle and colleagues [16] (“Keadle Women’s Health VM (2014)” option in ActiLife 6). Because most algorithms were developed from data that was derived through waist-worn devices, a correction of the wrist-derived data is necessary. This can be done by the “worn on wrist” option within the ActiLife 6 software package which is based on Actigraphs internal research and development (https://actigraphcorp.my.site.com/support/s/article/What-does-the-Worn-on-Wrist-option-do-in-the-Scoring-tab). According to personal communication with the manufacturer, this correction does not affect the step count calculation. The algorithm used to measure step counts is proprietary information and therefore not publicly available.

### Actigraphy Data Cleaning: Activity Variables

Data cleaning of the processed actigraphy data was done with the R software package, version 3.6.2 [20]. The first and last day of each individual’s wear time was excluded, because it did not contain information for the whole day and presumably was heavily confounded by measurements that were recorded during the time the device was delivered to or from the study site. Daily wear time was calculated by adding up the wear time periods for each individual day. Average daily wear time was calculated by dividing the cumulative wear time (over the whole wear time period) by the number of days the monitor was worn. Only participants who wore the device for 5 days or more consecutively with an average daily wear time of 15 hours or more were included in the statistical analyses presented in this study.

### Actigraphy Data Cleaning: Sleep Variables

Data on sleep time was available for 784 participants. We excluded 39 participants because they did not fulfill our pre-defined wear time requirements. One participant was excluded because he provided data on less than three sleep time periods. Time in bed and sleep time were extracted from data obtained through the GT3X-monitor by the algorithms implemented in the ActiLife 6 software package. The monitor was handed out and collected directly at the study center, all available data on sleep time is assumed to be valid. Therefore, all available data on sleep time and time in bed was used in this study.

The average sleep time at night was calculated by dividing the sum of all recorded sleep time periods by the number of sleep time periods. If the number of sleep time periods exceeded the number of nights (assessed as number of days the monitor was worn plus one), the discrepancy needs to be attributed to longer wake periods during the night or sleep time during the day (in addition to sleep time at night). In these cases, the average sleep time per night was calculated as follows: av. sleep time = sum of all sleep time periods/number of days monitor was worn +1.

A subgroup of participants that partly overlaps with participants that wore the GT3X-monitor filled in the Pittsburgh Sleep Quality Index sleep questionnaire (PSQI). Participants whose difference between the objective (GT3X-monitor) and self-reported (PSQI questionnaire) sleep time exceeded 50% of the self-reported sleep time, were excluded, because a measurement error seems to be very likely (n=24). After exclusion, sleep time variables obtained from the GT3X-monitor and ActiLife 6 were available for 720 participants. A subgroup of 410 participants provided additionally self-reported information about the sleep time.

### Rapid Assessment of Physical Activity (Questionnaire)

Self-reported PA was assessed via the Rapid Assessment of Physical Activity (RAPA) [21]. RAPA consists of seven questions about frequency and intensity of PA throughout the week. It is scored by choosing the highest ranked affirmed question which leads to classification in groups between 1 (sedentary) and 7 (active).

### Clinical Phenotypes and blood parameters

Age and sex were documented during 1:1-interviews with trained study personnel. Height and weight were measured with the electronic measuring station “seca 763” (SECA, Germany). All blood parameters were measured in a standard laboratory. Cholesterol, triglycerides, high density lipoprotein cholesterol (HDL-C) and low density lipoprotein cholesterol (LDL-C) were determined by enzymatic colorimetry. Serum fasting glucose was measured using the UV test. A turbidimetric inhibition immunoassay (TINIA) was employed to quantify glycated hemoglobin A1c.

### Imputation of Missing Values in actigraphy variables

To be able to make use of the accelerometric variables as covariates in further analyses that may e.g. include the complete number of participants (n=1100), missing values were imputed. The substitution of missings by the variable mean attenuates possible correlations to other variables, which is known to cause problems in multivariate analyses for which we want to use it in the future. Hence, we chose to employ stochastic regression imputation, which, in contrast to deterministic regression imputation, includes a random error term to predict the value of the missing data points based on variables known for all participants (chronological age, sex, BMI). Results of imputation were manually checked for plausibility. Of the 1,100 participants that were included in the analyzed cohort, 1,098 participants provided information about their BMI, age and sex. To be able to include as many participants as possible for the imputation, less strict inclusion criteria were employed. In concordance with previous analyses [22], participants who provided data on two days or more with an average wear time of ten hours or more were included. Missing values for 306 participants (37.9%) were substituted by stochastic regression imputation with R’s “mice” package [23].

### Statistical Analyses

The statistical tests were done with the R software package, version 3.6.2 [20]. Figures were produced with R’s “ggplot2” package [24]. A p-value <0.05 was used to define statistical significance. Difference between group’s mean was assessed with t-tests and analyses of variance (ANOVA). The correlation plot was drawn with the *ggcorr* function of the “ggplot2” extension “GGally”. We performed available cases analyses; therefore, participants were only excluded from an analysis if they did not provide data on one of the variables included in the analysis. The number of observations is stated for every analysis. Please note that the imputed values described above were only used for analyses shown in Supplementary Table 2.

## Results

### Cohort Characteristics and Wear Time

Descriptive statistics are displayed in Table 1. Of all 797 participants who wore the accelerometric monitor, 760 fulfilled our wear-time based inclusion criteria. Participants had a mean age of 75.6 years (SD: 3.8 years, range 66.0 – 94.1 years) and 53% of the analyzed sample were women. Participants wore the device for on average six days for on average 23.2 hours (SD: 1.3 hours) per day (Table 1). To identify potential under- or overestimation of total PA due to potential wear time preferences over the day, the week, or months of the year, stratified wear time validation analyses were conducted and no systematic irregularities in wear time were found (Figure 1 and Supplementary Figure 1). Furthermore, no variance in wear time with respect to the number of days the device was already worn was found (data not shown).

**Table 1:** Cohort characteristic of BASE-II participants that provided accelerometric data obtained through ActiGraph’s GT3X monitor.

| Variables |  | n | mean, % | sd | min | max |
| --- | --- | --- | --- | --- | --- | --- |
| Age (years) |  | 760 | 75.61 | 3.81 | 65.95 | 94.07 |
| Sex | Male | 357 | 46.97 |  |  |  |
|  | Female | 403 | 53.03 |  |  |  |
| RAPA Score | 1 | 2 | 0.26 |  |  |  |
|  | 2 | 4 | 0.53 |  |  |  |
|  | 3 | 103 | 13.57 |  |  |  |
|  | 4 | 272 | 35.84 |  |  |  |
|  | 5 | 91 | 11.99 |  |  |  |
|  | 6 | 226 | 29.78 |  |  |  |
|  | 7 | 61 | 8.04 |  |  |  |
| Seldom/never physically active<sup>1</sup> | Yes | 96 | 12.65 |  |  |  |
|  | No | 663 | 87.35 |  |  |  |
| Vector Magnitude (counts) |  | 760 | 19,310,000.00 | 5,481,000.00 | 4,604,000.00 | 43,830,000.00 |
| Step Count |  | 760 | 12,050.00 | 3,415.00 | 2,664.00 | 26,070.00 |
| Energy Expenditure (kcal) |  | 760 | 1,540.00 | 574.40 | 45.72 | 4,734.00 |
| MET (average/day) |  | 760 | 1.42 | 0.16 | 1.03 | 2.01 |
| Physical Activity: Sedentary (%/hour) |  | 760 | 49 | 7 | 26 | 73 |
| Physical Activity: Light (%/hour) |  | 760 | 29 | 5 | 14 | 49 |
| Physical Activity: Moderate (%/hour) |  | 760 | 22 | 7 | 2 | 47 |
| Average Wear Time (hours/day) |  | 760 | 23.15 | 1.31 | 15.59 | 24.00 |
| Sleep time (hours/night), GT3X-Monitor |  | 720 | 6.91 | 1.25 | 3.27 | 11.49 |
| Sleep time (hours/night), questionnaire |  | 410 | 7.07 | 1.15 | 4 | 11 |
| Difference sleep time (GT3X – questionnaire) |  | 410 | -0.08 | 1.37 | -4.45 | 3.32 |

<sup>1</sup>According to RAPA item 1

**Figure 1:**
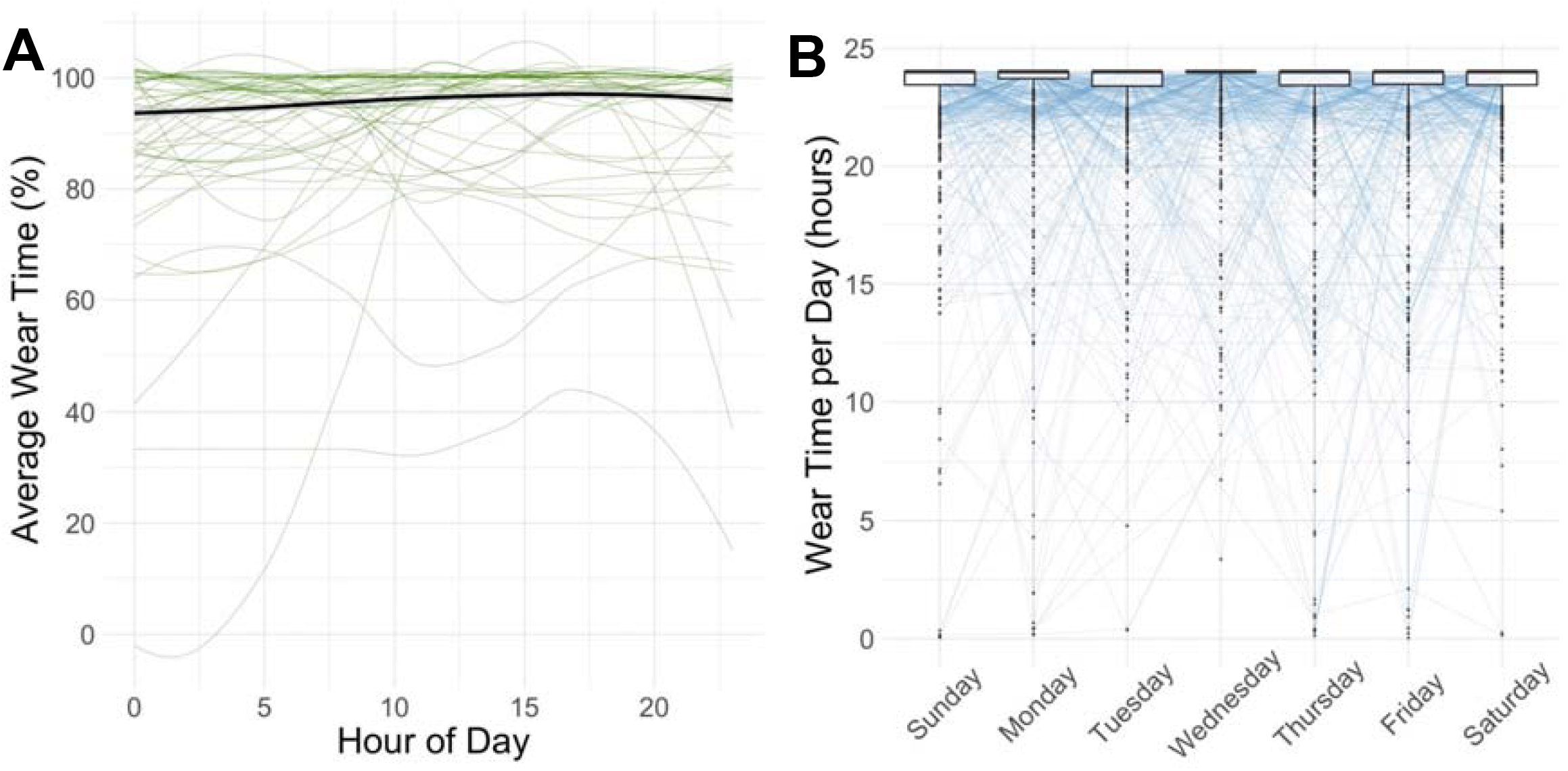
Analyses of average wear time (A) as a function of hour of the day and boxplots of average daily wear time (B) stratified by day of the week. Consistently high wear time was present through the day and the week. Median, hinges (25th and 75th percentile) and Tukey-style whiskers (1.5* inter-quartile-range) are displayed in the boxplots. Boxplot outliers were defined as values more extreme than 1.5*IQR.

### Physical Activity

Descriptive statistics of the PA variables that were derived from the GT3X monitor are shown in Table 1. We found higher PA in men when assessed as Vector Magnitude, Step Count, Energy Expenditure and cut-off defined PA (sedentary, light, and moderate, p<=0.002, Supplementary Table 1).

A moderate to high positive correlation was found between Vector Magnitude, Step Counts, Energy Expenditure, time spent in moderate PA and Metabolic Equivalent of Task (MET, Pearson’s r between 0.5 and 0.97). The highest correlation was found between MET and Energy Expenditure (Pearson’s r= 0.93) and between “moderate” PA (which indicates the time spent in an average hour in the most intense PA) and Vector Magnitude (Pearson’s r=0.97, Figure 2).

**Figure 2:**
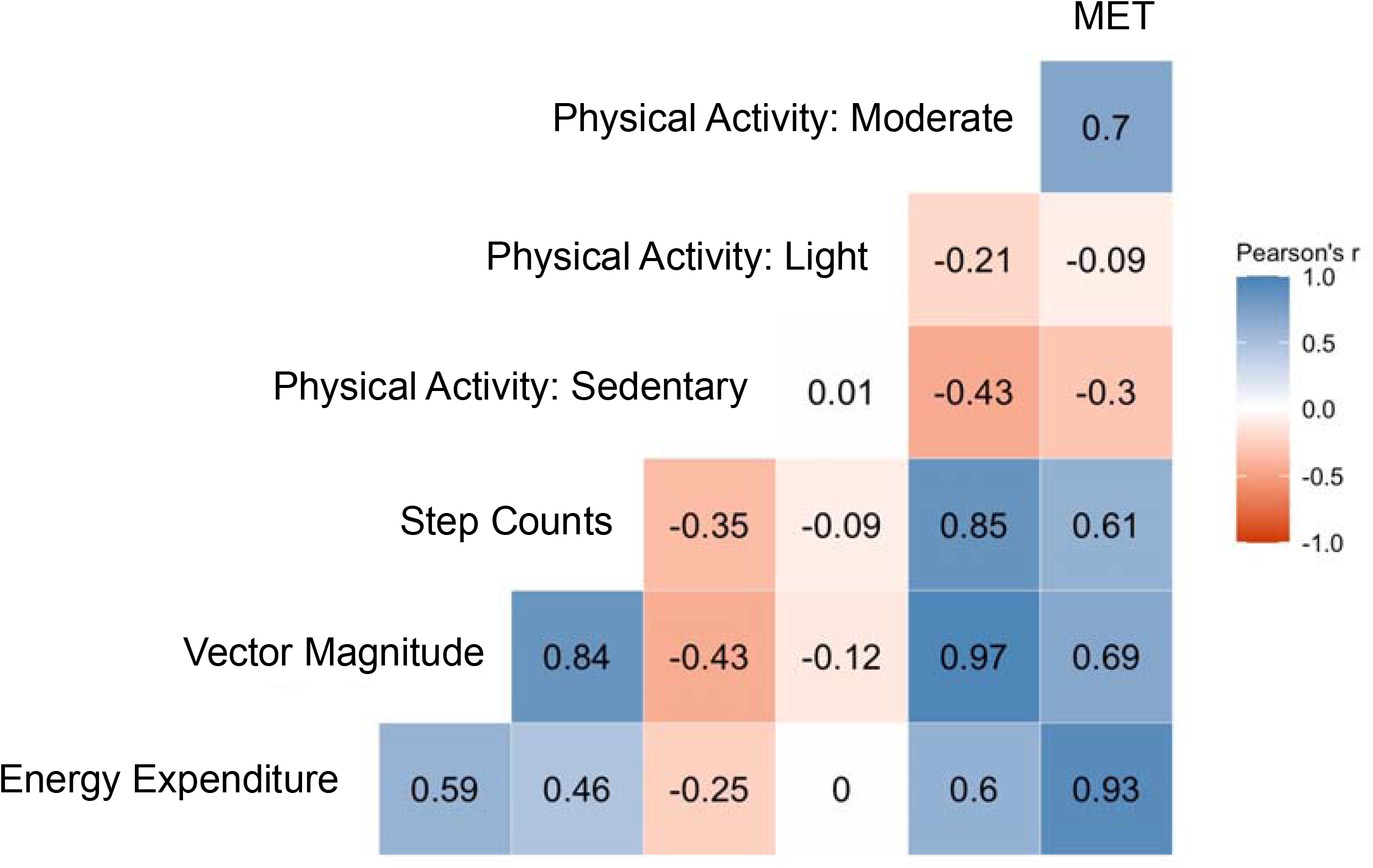
Heatmap of Pearson’s correlation between the available accelerometric variables. Correlation coefficients were calculated as Pearson’s r.

As expected, the two variables indicating lower PA - sedentary and light PA-were negatively associated with the other PA variables (Pearson’s r between 0 and −0.4, Figure 2).

Normalized values of the different activity variables over the course of the day are depicted in Supplementary Figure 2. The available variables reflect the expected daily activity profile. We found only moderate seasonal changes in activity over the course of the year (Supplementary Figure 3) with the highest amount of PA around April. The differences are, however, rather small, and we would not expect them to impose considerable bias to our analyses.

### Comparison of objective and subjective (self-rated) physical activity

To further explore how objective and subjective PA were linked in this cohort, we compared results obtained from the accelerometer with self-reported PA assessed with the well-established Rapid Assessment of Physical Activity (RAPA). The RAPA contains seven yes/no-questions and classifies participants based on self-rated PA. We found significant differences between objectively assessed PA indicators and the RAPA groups on all accelerometric measures except for Energy Expenditure and average minutes per hour spent with light PA (p<=0.02, ANOVA, Figure 3).

**Figure 3:**
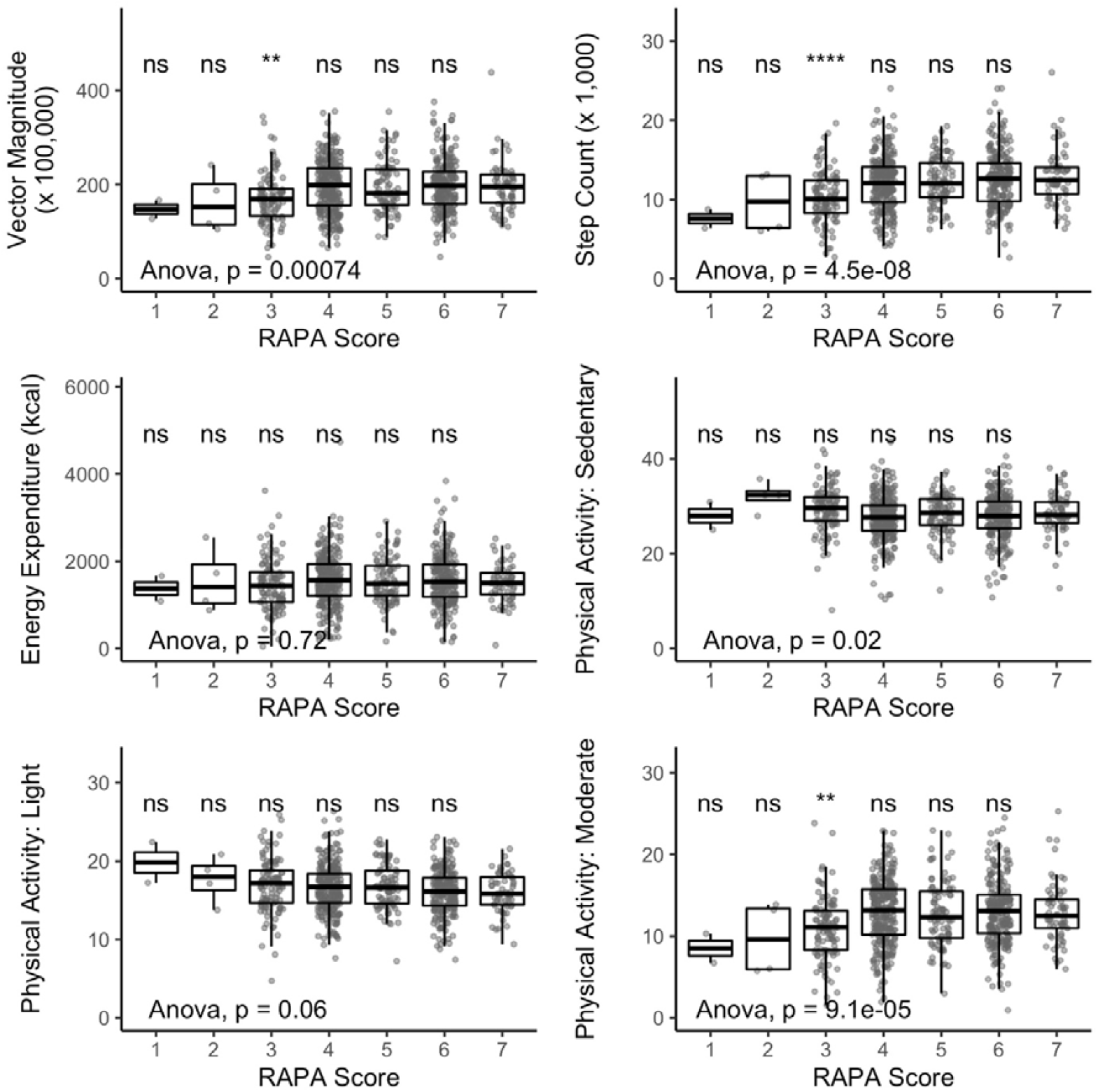
Results in activity variables stratified by self-rated physical activity (assessed via RAPA questionnaire). Statistical significance of difference between means was assessed by ANOVA. Difference between the highest self-rate activity (RAPA = 7) and the other RAPA groups were tested for statistical significance with t-test. N=759. **p<=0.01, ****p<=0.0001

However, when performing a post-hoc analysis comparing the highest PA group (RAPA group 7 = “active”) with the other RAPA groups, significant differences were only found in group 3 in Vector Magnitude, Step Count, and moderate PA (t-Test, Figure 3). Furthermore, we found differences in accelerometric variables to be higher between RAPA groups indicating low self-rated PA (e.g. group 1 to 3, Figure 3) compared to differences between RAPA groups indicating that the participants were “under-active” or “active” with respect to PA (groups 4 to 7). Sex-stratified analyses showed similar results and are displayed in Supplementary Figure 4.

Subsequently, we examined differences in the accelerometric variables between participants who rated themselves based on the first RAPA question to be “seldom/never active” or “active”. We found that participants who considered themselves as physically active showed higher PA levels measured as Vector Magnitude, step counts, and cut-off defined moderate PA compared to the more sedentary participants stating that they were seldom or never active (p<0.05, t-Test, Figure 4). In sex-stratified follow-up analyses, this association remained significant for step counts only in the female and male subgroup and for moderate PA in the female subgroup only (Supplementary Figure 5).

**Figure 4:**
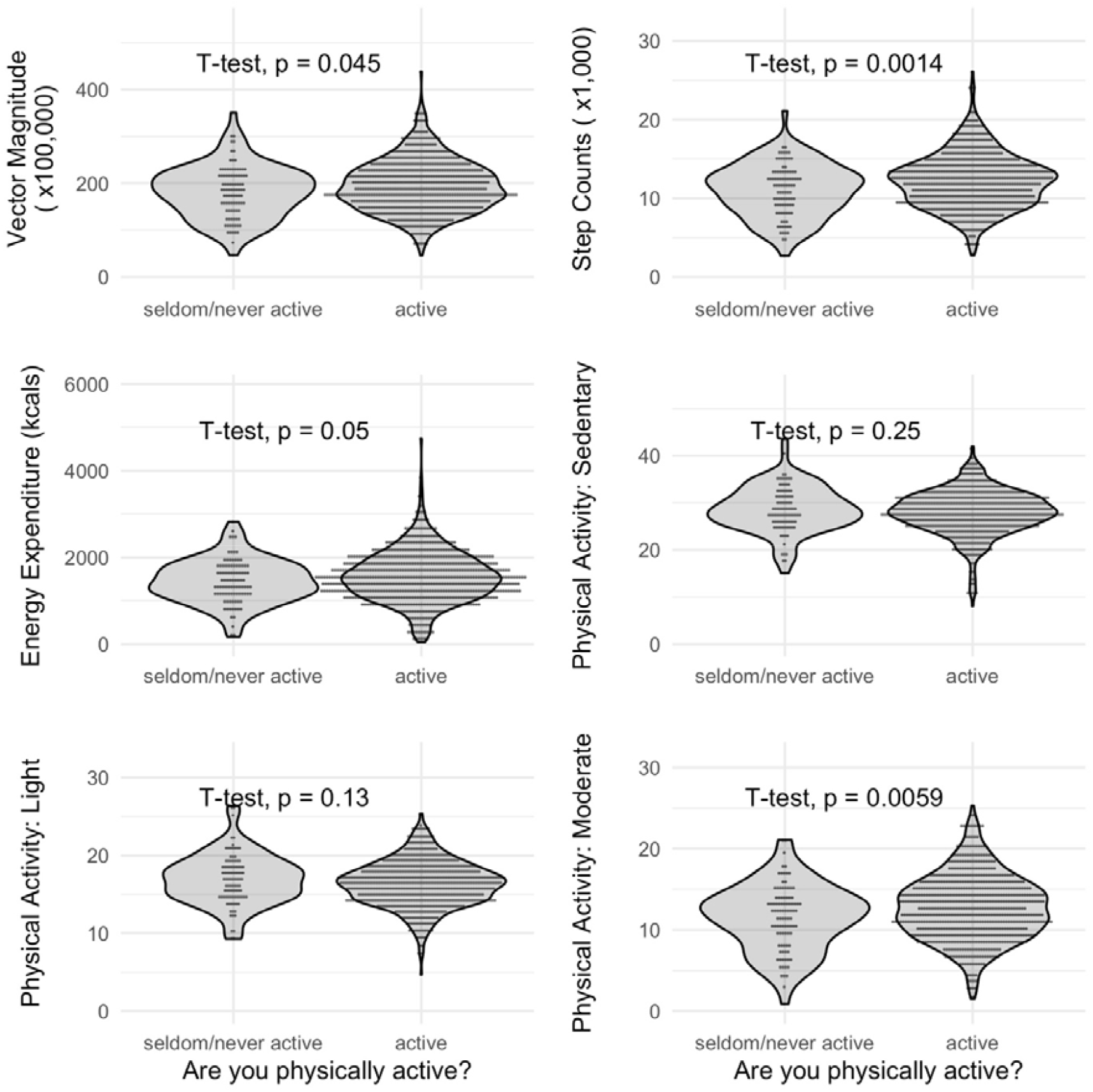
Violin plots of accelerometric activity variables of participants that reported to be seldom/never active or to be active. Statistical significance of difference between means was calculated using t-test. N=759.

### Relationship between physical activity, age, BMI, and known activity-associated blood parameters

We found a weak negative correlation between age and PA (Vector Magnitude, step count, energy expenditure, MET, and moderate PA; Pearson’s r between −0.16 and −0.18, n=760, Table 2). No significant association was found between age and variables indicating sedentary or light PA (Pearson’s r<0.1, Table 2). Interestingly, a moderate positive correlation was found between energy expenditure and BMI (Pearson’s r=0.4), although step counts and vector magnitude showed a negative correlation with this variable (Pearson’s r=-0.2, n=760, Table 2). This finding most likely results from the way energy expenditure is calculated by the Actilife software package which incorporates the individual’s weight. Weak correlations were found between activity variables and TC, LDL-C, TG, fasting glucose and HbA1c. The strongest correlation was found between HDL and vector magnitude, step count and cut-off defined moderate PA (Pearson’s r= between 0.21 and 0.25, n=750, Table 2).

**Table 2:**
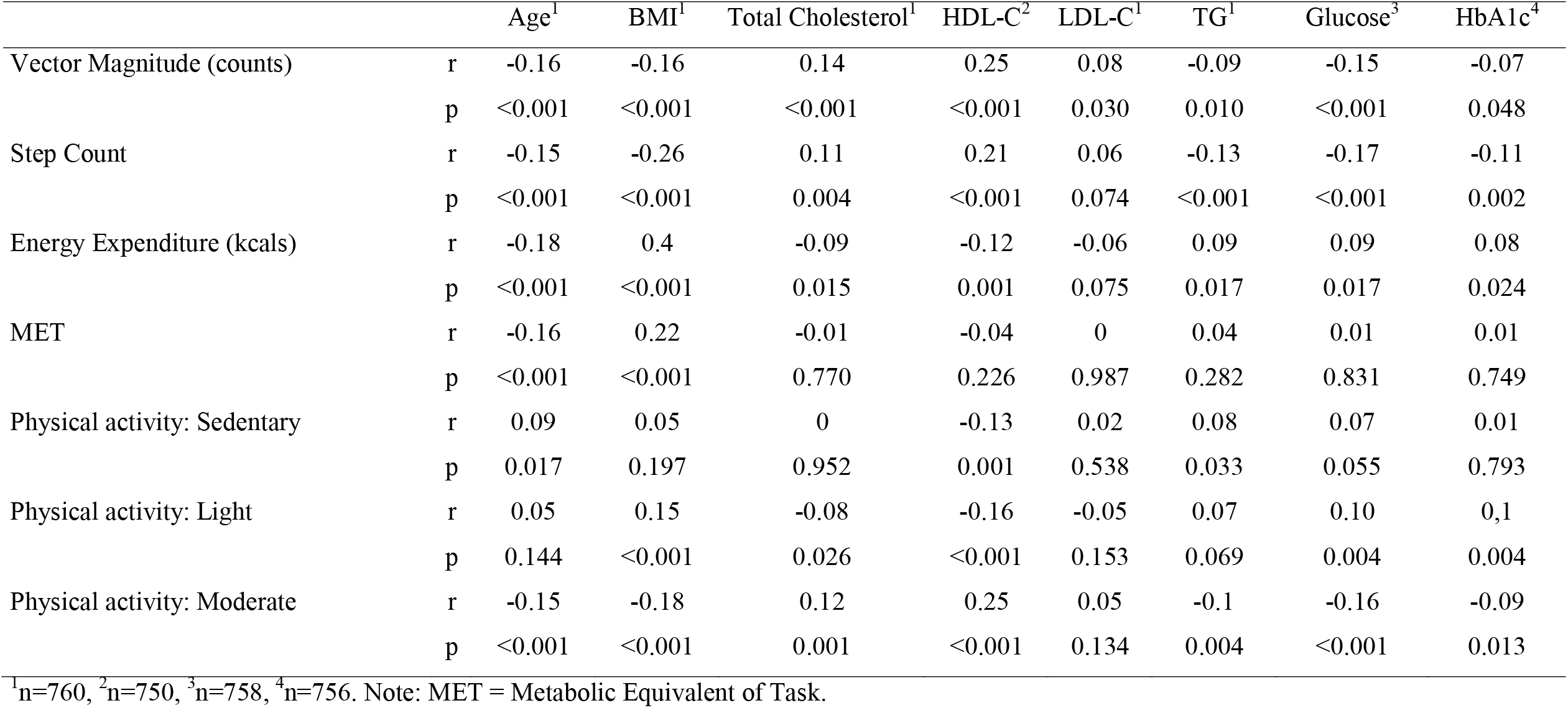
Table of correlation between activity variables (Actigraph), chronological age, BMI and blood parameters. Correlation was calculated as Pearson’s r.

|  |  | Age<sup>1</sup> | BMI<sup>1</sup> | Total Cholesterol<sup>1</sup> | HDL-C<sup>2</sup> | LDL-C<sup>1</sup> | TG<sup>1</sup> | Glucose<sup>3</sup> | HbA1c<sup>4</sup> |
| --- | --- | --- | --- | --- | --- | --- | --- | --- | --- |
| Vector Magnitude (counts) | r | -0.16 | -0.16 | 0.14 | 0.25 | 0.08 | -0.09 | -0.15 | -0.07 |
|  | p | <0.001 | <0.001 | <0.001 | <0.001 | 0.030 | 0.010 | <0.001 | 0.048 |
| Step Count | r | -0.15 | -0.26 | 0.11 | 0.21 | 0.06 | -0.13 | -0.17 | -0.11 |
|  | p | <0.001 | <0.001 | 0.004 | <0.001 | 0.074 | <0.001 | <0.001 | 0.002 |
| Energy Expenditure (kcal) | r | -0.18 | 0.4 | -0.09 | -0.12 | -0.06 | 0.09 | 0.09 | 0.08 |
|  | p | <0.001 | <0.001 | 0.015 | 0.001 | 0.075 | 0.017 | 0.017 | 0.024 |
| MET | r | -0.16 | 0.22 | -0.01 | -0.04 | 0 | 0.04 | 0.01 | 0.01 |
|  | p | <0.001 | <0.001 | 0.770 | 0.226 | 0.987 | 0.282 | 0.831 | 0.749 |
| Physical activity: Sedentary | r | 0.09 | 0.05 | 0 | -0.13 | 0.02 | 0.08 | 0.07 | 0.01 |
|  | p | 0.017 | 0.197 | 0.952 | 0.001 | 0.538 | 0.033 | 0.055 | 0.793 |
| Physical activity: Light | r | 0.05 | 0.15 | -0.08 | -0.16 | -0.05 | 0.07 | 0.10 | 0.1 |
|  | p | 0.144 | <0.001 | 0.026 | <0.001 | 0.153 | 0.069 | 0.004 | 0.004 |
| Physical activity: Moderate | r | -0.15 | -0.18 | 0.12 | 0.25 | 0.05 | -0.1 | -0.16 | -0.09 |
|  | p | <0.001 | <0.001 | 0.001 | <0.001 | 0.134 | 0.004 | <0.001 | 0.013 |

<sup>1</sup>n=760, <sup>2</sup>n=750, <sup>3</sup>n=758, <sup>4</sup>n=756. Note: MET = Metabolic Equivalent of Task.

### Imputation of Incomplete Cases

To achieve a complete dataset for future analyses, stochastic regression imputation was used to substitute missing values of 306 participants as a function of chronological age, sex, and BMI. To validate the resulting dataset, the correlation analyses described above were repeated in the extended dataset (Supplementary Table 2). As expected and due to the methodological approach chosen for the imputation, correlation with age and BMI in the extended dataset was very similar to the results found in original dataset that includes only the measured results. However, a very high degree of agreement in correlation between variables was found for the blood parameters, as well (Supplementary Table 2).

### Sleep

Self-reported data about sleep time was available for a subgroup 410 participants of all participants who wore the accelerometric monitor. An average sleep time of 7.1 hours (SD=1.15 hours, range: 4 – 11 hours, n=410, Table 1) was reported by the participants in the PSQI questionnaire. Very similar results were measured with the GT3X monitor (mean sleep time = 6.91 hours, SD= 1.3 hours, range: 3.3 – 11.5 hours, n=720, Table 1). The average difference between objective and subjective sleep time was −0.1 hours (SD= 1.4, range: −4.5 – 3.3 hours, n=410, Table 1).

Subjective and objective sleep time were moderately correlated (Pearson’s r=0.31, p>0.001, n=410, Supplementary Figure 6 A). This was also true for sex-stratified subgroup analyses (women: Pearson’s r=0.34, p<0.001 and men: Pearson’s r=0.29, p<0.001, Supplementary Figure 6 B).

## Discussion

In this study we analyzed subjective and objective assessments of PA from 760 participants originating from the BASE-II study aged between 66 and 95 years (53.0% women) who wore an accelerometer for on average six consecutive days and completed the RAPA questionnaire. Wear time was found to be high and independent from the time of the day, weekday, month, or season. Expected correlations were found between objectively assessed activity levels and self-reported PA, BMI and blood parameters known to be associated with PA.

Step counts in this cohort were in a comparable range with results from cohorts with a similar age distribution and accelerometric data derived from wrist-worn accelerometers [9, 11]. Mandigout and colleagues measured 11,060 steps per day in 22 participants (mean age (SD): 76.6 (4.7), 36% female) from a wrist worn device in contrast to 5,922 steps that were obtained from waist worn monitors in the same participants at the same time [11]. A similar degree of overestimation was found by Kamada and colleagues who examined 94 women (mean age (SD): 71.9 (6.0) years) and reported 10,107 (wrist worn device) and 5378 steps (waist worn device) for the same activity period. Both authors conclude that the step count variable by the GT3X monitor (respectively the ActiLife software) is vastly overestimated when obtained from wrist-worn monitors. Although participants in the current study wore the accelerometer on their wrist without any comparison to waist-worn monitors, it is highly likely that a comparable degree of overestimation can be expected in our data, as well. Systematic overestimation may be attributed to the underlying proprietary ActiLife algorithm which was trained on data obtained from waist worn devices [9]. Furthermore, a waist worn device is closer to the body’s center of mass and might therefore be able to better capture the whole body’s acceleration [11]. Nevertheless, many sitting or standing activities may involve movements of arms. Thus, a wrist mounted accelerometer may detect this kind of acceleration induced by movements of the arms only, even when the participant is not walking [11]. It was argued that this measurement “may, however, be a good indicator of a more global amount of PA [physical activity]” [11]. Additionally, it is important to mention that the step count variable obtained from wrist and waist worn monitors were correlated in a previous study [11] and correlate substantially when classifying participants’ activity in quintiles [9]. Similar results were reported for Vector Magnitude [9, 10]. These findings suggest that although the activity variables obtained from wrist worn devices probably cannot be used as absolute values, they can be informative on differences in PA within the analyzed group and might provide an even more complete assessment of PA as the waist worn monitors because it is able to include upper limb activity.

Our findings support this assumption showing that activity variables and BMI as well as PA and blood parameters are associated. For instance, participants with a higher BMI tend to have recorded less steps per day (Pearson’s r = −0.3). Interestingly, a positive correlation between BMI and energy expenditure was found (Pearson’s r = 0.4). This inverse finding, however, probably needs to be attributed to the underlying algorithm for energy expenditure, which incorporates body weight and acknowledges the higher amount of energy that would be needed to carry a larger body mass. Additionally, weak correlations were found between higher HDL-C and higher levels of activity measured as Vector Magnitude, step counts, and time spent with moderate PA (Pearson’s r between 0.21 and 0.25). Correlations between blood parameters and Vector Magnitude and step counts found in this study were consistent with previous results reported by Wolff-Hughes and colleagues for TC, LDL-C, HDL-C, TG, glucose, and HbA1c [25], although the effect size of the association was difficult to compare due to methodological differences. Additionally, the direction of associations was the same for the bout-defined measure of moderate PA with the only exception of TC and LDL-C [25]. We would like to point out several limitations to this study. First, due to the position the monitor was worn, we were not able to measure accurately the absolute number of step taken. However, this phenomenon is well-known and does not hinder the use of this variable relatively within the analyzed group. Second, the measured data was assessed under every-day life conditions and no diary-based information on PA was available for this period. Therefore, we were not able to directly validate the objective data with information acquired under laboratory conditions or through detailed records of activity. However, we were able to show that the accelerometric activity data showed expected associations with age, BMI and fitness-associated blood parameters. Therefore, and because the employed monitor is one of the most frequently used accelerometers in epidemiologic studies, we are confident that the measured data provides valuable information about the participant’s individual PA level. Third, only a subgroup of the total sample of 1,100 participants of the analyzed cohort provided accelerometric data. To be able to make use of the whole dataset in future analyses, the missing values were substituted by a stochastic regression imputation. Although the fraction of imputed values in the final dataset is large (27.9%), results of correlation analyses between the extended dataset and blood parameters make the values seem plausible and therefore potentially valuable for use in subsequent analyses.

Strengths of this study are the large number of participants who provided accelerometric data. Additionally, the wear time of the accelerometers was very high and independent from time of day, day of week or season. Due to the extensive and rich dataset of BASE-II we were able to relate the objectively measured PA data as well as sleep time to subjectively reported PA levels and self-reported sleep time. Additionally, we examined the relationship between the accelerometric variables and fitness-associated blood parameters. The different PA variables derived from the accelerometer can be used to comprehensively examine different aspects of PA in future analyses of BASE-II data e.g. in the context of cardio-metabolic health.

In conclusion, activity variables measured with the GT3X monitor and processed with the ActiLife 6 software package provided valid information on different aspects of PA for 760 participants in the Berlin Aging Study II.

## Data Availability

Due to concerns for participant privacy, data are available only upon reasonable request. Please contact Ludmila Mueller, scientific coordinator, at, for additional information.

## Author contributions

Conceived and designed the study: ID, SD, VMV. Analyzed the data: DÖ and VMV. Involved in first steps of data analyses: JK. Supervised the project: ID. Wrote the manuscript: VMV. All authors revised and approved the manuscript.

## Data availability statement

Due to concerns for participant privacy, data are available only upon reasonable request. Please contact Ludmila Müller, scientific coordinator, at, for additional information.

