## Supplementary Material for "Subjective and Objective Assessment of Physical Activity in Older Adults: Results from Berlin Aging Study II"

Charité - Universitätsmedizin Berlin

Lipid Clinic at the Interdisciplinary Metabolism Center,

Biology of Aging Group

Augustenburger Platz 1

13353 Berlin

Supplementary Material:

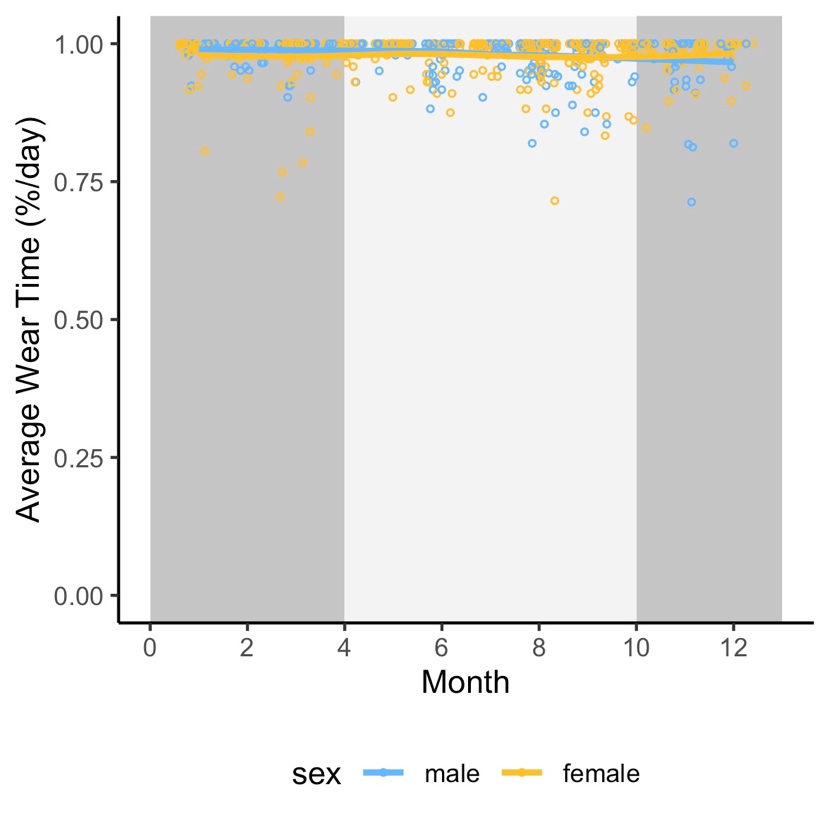

**Supplementary Figure 1: Average individual wear time of each participant per month of the first day of wear time.** Season are indicated by background color (dark grey = winter, light grey = summer).

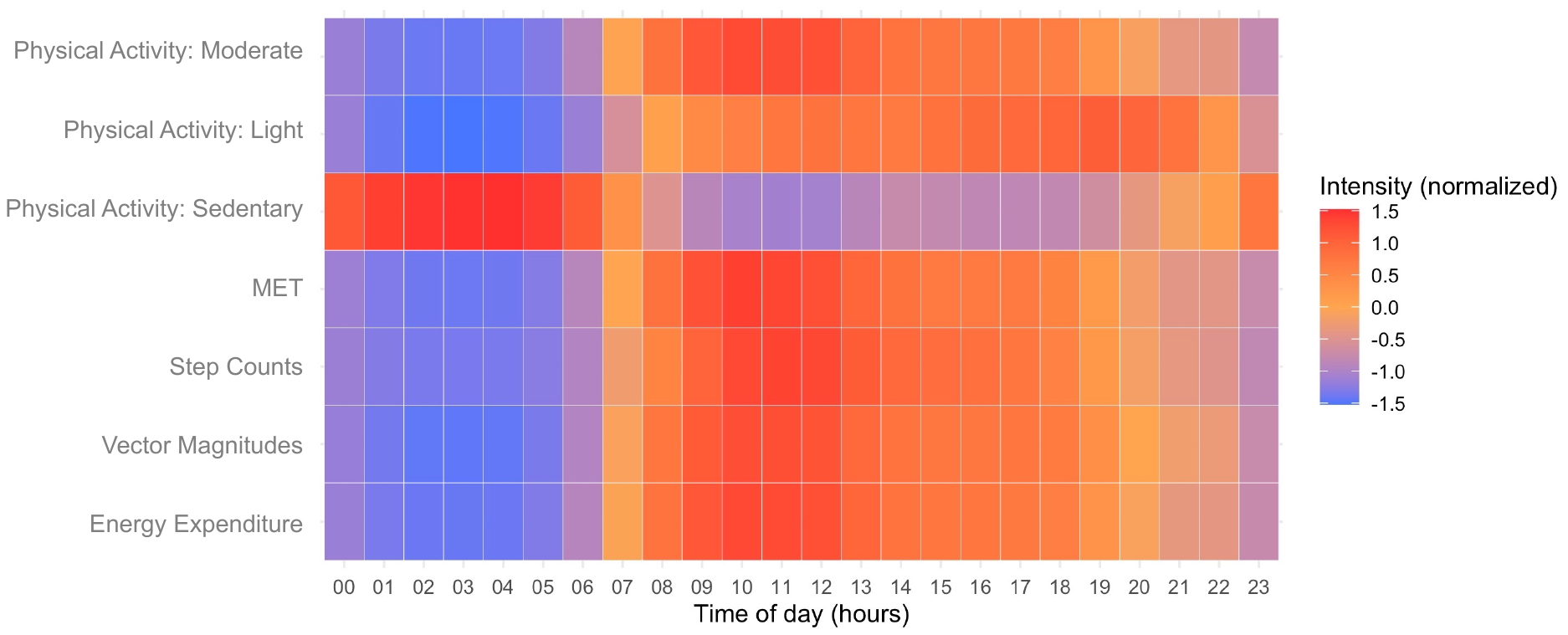

**Supplementary Figure 2: Heatmap of the normalized average values of the available activity variables over the course of the day.**

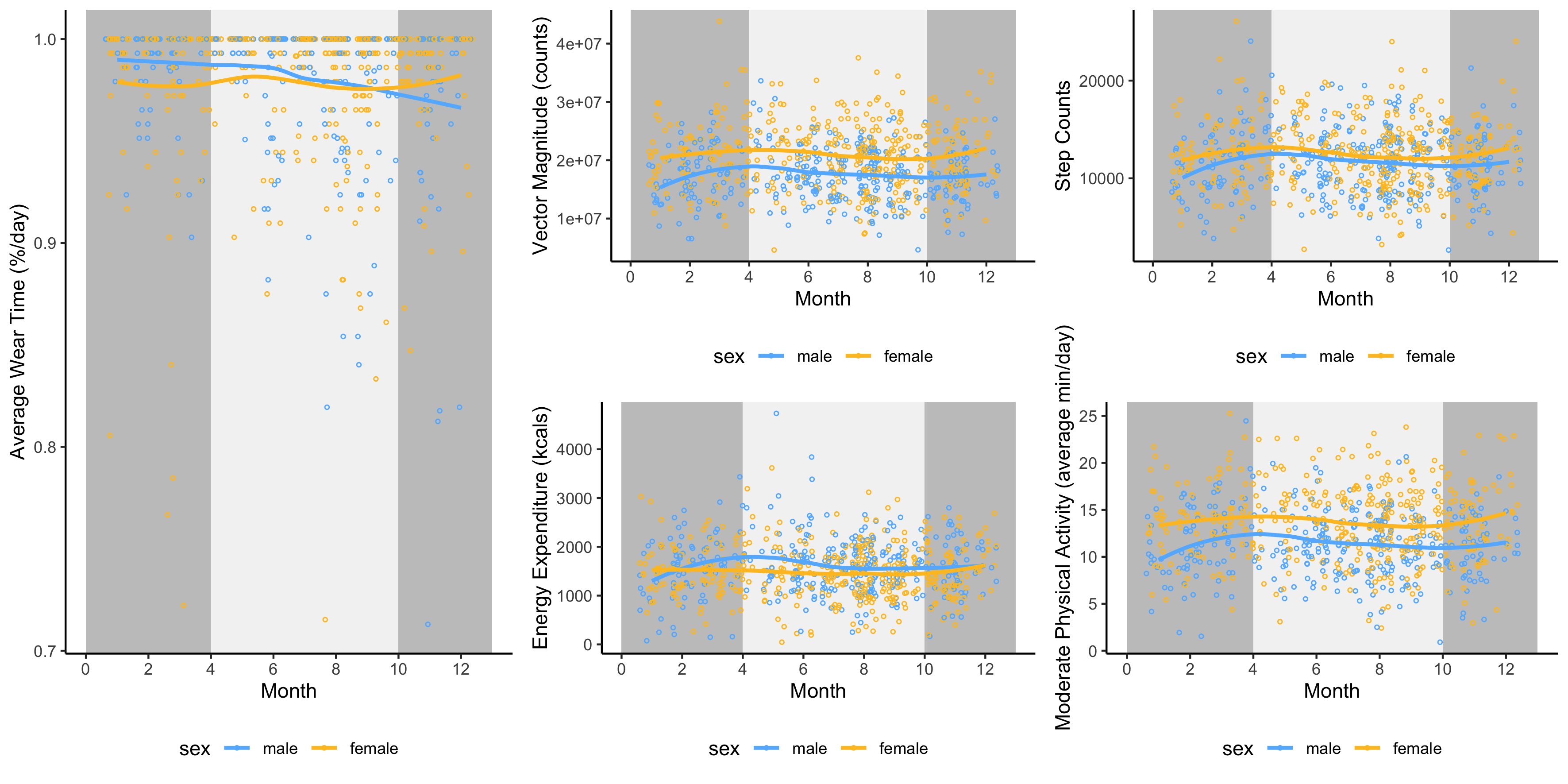

**Supplementary Figure 3: Average physical activity of BASE-II participants over the course of the year.** Season is indicated by background color (dark grey = winter, light grey = summer).

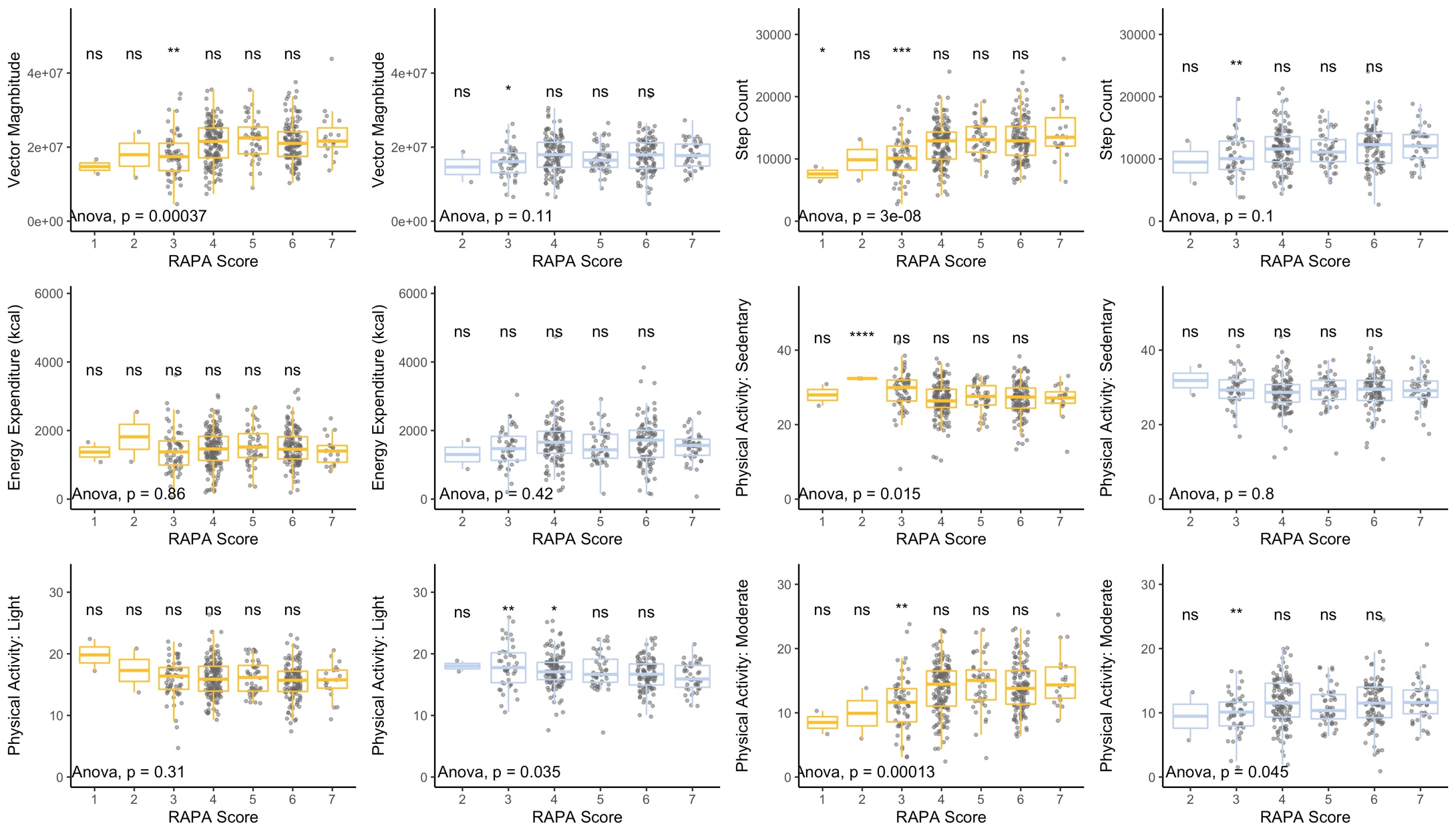

**Supplementary Figure 4: Results in activity variables stratified by self-rated physical activity (assessed via RAPA questionnaire) and sex (female = yellow, male = blue).** Statistical significance of difference between means was assessed by ANOVA. Difference between the highest self-reported activity (RAPA = 7) and the other RAPA groups were tested for statistical significance with t-test.

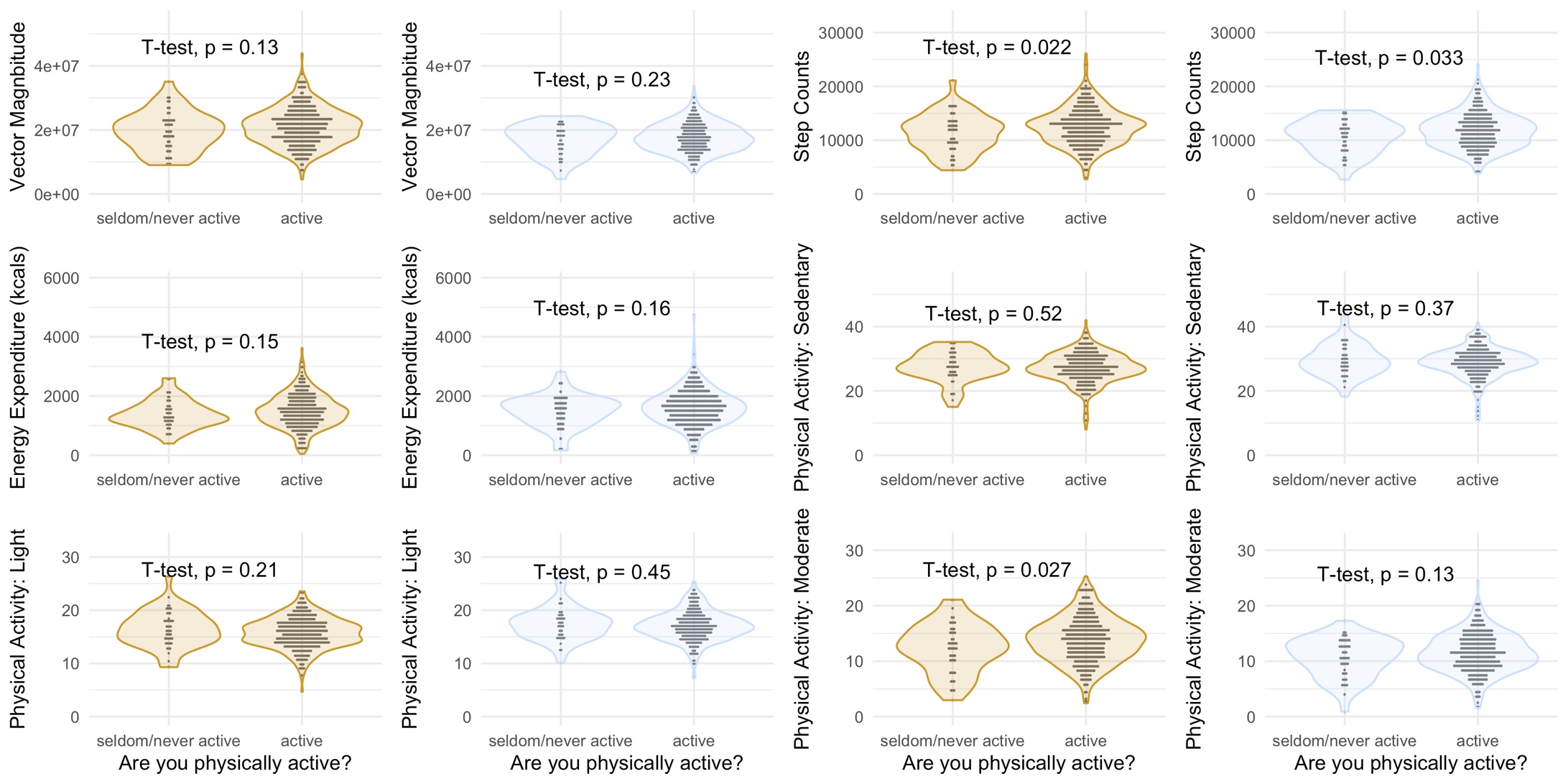

**Supplementary Figure 5: Sex-stratified violin plots of accelerometric activity variables of participants that reported to be seldom/never active or to be active (female = yellow, male = blue).** Statistical significance of differences was assessed by t-test.

**A**
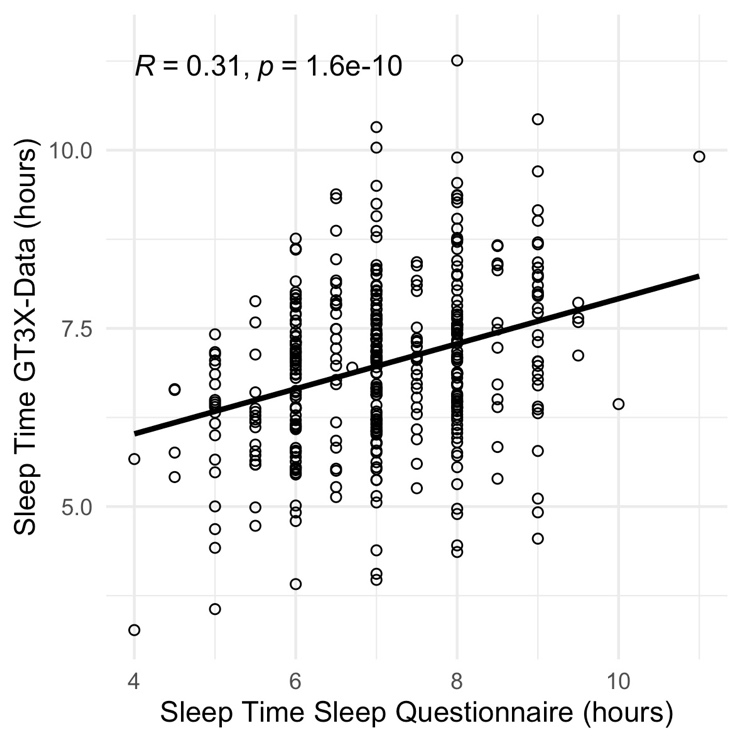
 **B**
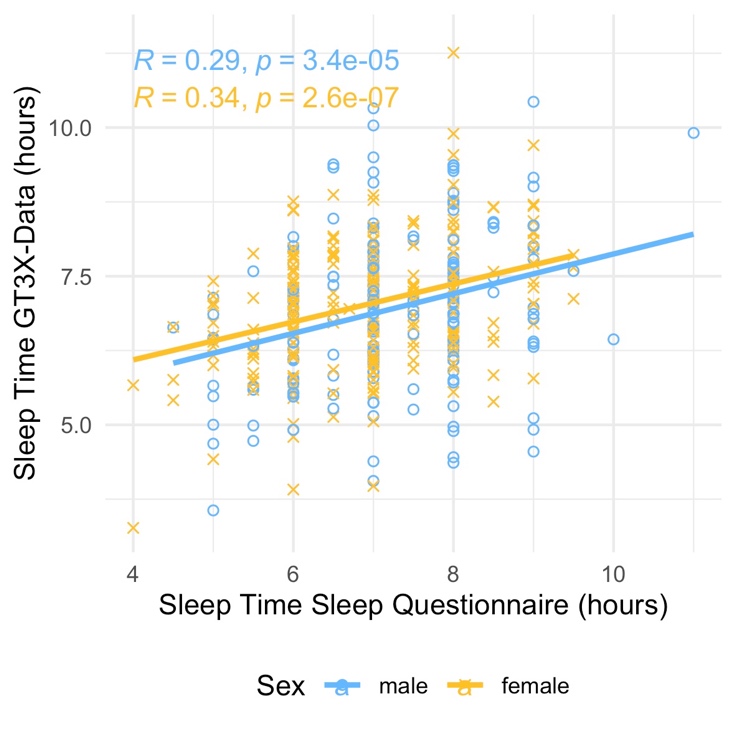

**Supplementary Figure 6: Scatterplots of sleep time assessed with the GT3X monitor and questionnaire based sleep time.** A) All participants were included (n=410) and B) sex-stratified subgroup analyses were conducted.

**Supplementary Table 1: Sex-stratified cohort characteristics**

|  |  |  | Women | | | | |  | Men | | | | |  |  |
| --- | --- | --- | --- | --- | --- | --- | --- | --- | --- | --- | --- | --- | --- | --- | --- |
|  |  |  | n | mean | sd | min | max |  | n | mean | sd | min | max |  | p-value |
| Age (years) |  |  | 403 | 75.75 | 3.55 | 67.13 | 94.07 |  | 357 | 75.46 | 4.08 | 65.95 | 90.03 |  | 0.295 |
| Sex | Male |  |  |  |  |  |  |  |  |  |  |  |  |  |  |
|  | Female |  |  |  |  |  |  |  |  |  |  |  |  |  |  |
| RAPA Score | 1 |  | 2 | 0.50 |  |  |  |  | 0 | 0.00 |  |  |  |  | 0.012 |
|  | 2 |  | 2 | 0.50 |  |  |  |  | 2 | 0.56 |  |  |  |  |  |
|  | 3 |  | 59 | 14.64 |  |  |  |  | 44 | 12.36 |  |  |  |  |  |
|  | 4 |  | 151 | 37.47 |  |  |  |  | 121 | 33.99 |  |  |  |  |  |
|  | 5 |  | 44 | 10.92 |  |  |  |  | 47 | 13.20 |  |  |  |  |  |
|  | 6 |  | 126 | 31.27 |  |  |  |  | 100 | 28.09 |  |  |  |  |  |
|  | 7 |  | 19 | 4.71 |  |  |  |  | 42 | 11.80 |  |  |  |  |  |
| Seldom/never physically active^1^ | yes |  | 49 | 12.16 |  |  |  |  | 47 | 13.20 |  |  |  |  | 0.747 |
|  | no |  | 354 | 87.84 |  |  |  |  | 309 | 86.80 |  |  |  |  |  |
| Vector Magnitude (counts) |  |  | 403 | 20,860,000.00 | 5,793,000.00 | 4,604,000.00 | 43,830,000.00 |  | 357 | 17,570,000.00 | 4,516,000.00 | 4,641,000.00 | 33,630,000.00 |  | <0.001 |
| Step Count |  |  | 403 | 12,430.00 | 3,510.00 | 2,740.00 | 26,070.00 |  | 357 | 11,620.00 | 3,255.00 | 2,664.00 | 24,040.00 |  | 0.001 |
| Energy Expenditure (kcals) |  |  | 403 | 1,480.00 | 552.70 | 45.72 | 3,617.00 |  | 357 | 1,606.00 | 591.60 | 74.54 | 4,734.00 |  | 0.002 |
| MET |  |  | 403 | 1.42 | 0.16 | 1.03 | 1.91 |  | 357 | 1.43 | 0.16 | 1.03 | 2.01 |  | 0.249 |
| Physical Activity: Sedentary (%/hour) |  |  | 403 | 48 | 7 | 26 | 70 |  | 357 | 50 | 7 | 28 | 73 |  | <0.001 |
| Physical Activity: Light (%/hour) |  |  | 403 | 28 | 5 | 14 | 49 |  | 357 | 30 | 4 | 20 | 45 |  | <0.001 |
| Physical Activity: Moderate (%/hour) |  |  | 403 | 24 | 7 | 5 | 47 |  | 357 | 20 | 6 | 2 | 42 |  | <0.001 |
| Average Wear Time (hours/day) |  |  | 403 | 23.09 | 1.35 | 15.59 | 24.00 |  | 357 | 23.22 | 1.26 | 16.19 | 24.00 |  | 0.157 |

**Supplementary Table 2: Correlation table of the whole dataset including imputed values for accelerometric variables. Correaltion was assessed as Pearson’s r.** Please note that the correlation coefficients for the subgroup with measured accelerometric data may vary from the results shown in the main manuscript because more strict inclusion criteria were applied to the dataset used for analyses that were presented in the main manuscript, as it focusses on the accelerometric data.

|  | BMI | | Age | | Cholesterol | | HDL | | LDL | | Triglycerides | | Glucose | | HbA1c | |
| --- | --- | --- | --- | --- | --- | --- | --- | --- | --- | --- | --- | --- | --- | --- | --- | --- |
|  | r | n | r | n | r | n | r | n | r | n | r | n | r | n | r | n |
| Energy Expenditure (kcals) | 0.39^**^ | 792 | -0.17^**^ | 792 | -0.09^*^ | 792 | -0.13^**^ | 782 | -0.06 | 792 | 0.07 | 792 | 0.08^*^ | 790 | 0.08^*^ | 788 |
| Energy Expenditure (kcals, imputed) | 0.39^**^ | 1098 | -0.16^**^ | 1098 | -0.06 | 1093 | -0.12^**^ | 1078 | -0.03 | 1091 | 0.07^*^ | 1092 | 0.09^*^ | 1090 | 0.09^*^ | 1089 |
| Vector Magnitude (counts) | -0.15^**^ | 792 | -0.15^**^ | 792 | 0.14^**^ | 792 | 0.24^**^ | 782 | 0.08^*^ | 792 | -0.11^*^ | 792 | -0.15^**^ | 790 | -0.07^*^ | 788 |
| Vector Magnitude (counts, imputed) | -0.15^**^ | 1098 | -0.14^**^ | 1098 | 0.17^**^ | 1093 | 0.22^**^ | 1078 | 0.12^**^ | 1091 | -0.07^*^ | 1092 | -0.14^**^ | 1090 | -0.06^*^ | 1089 |
| Step Count | -0.25^**^ | 792 | -0.14^**^ | 792 | 0.1^*^ | 792 | 0.2^**^ | 782 | 0.07^*^ | 792 | -0.14^**^ | 792 | -0.17^**^ | 790 | -0.11^*^ | 788 |
| Step Count (imputed) | -0.25^**^ | 1098 | -0.13^**^ | 1098 | 0.13^**^ | 1093 | 0.18^**^ | 1078 | 0.1^*^ | 1091 | -0.1^*^ | 1092 | -0.17^**^ | 1090 | -0.1^**^ | 1089 |
| Physical activity: Sedentary | 0.06 | 792 | 0.09 | 792 | -0.01 | 792 | -0.11^*^ | 782 | 0.02 | 792 | 0.01 | 792 | 0.04 | 790 | -0.01 | 788 |
| Physical activity: Sedentary (imputed) | 0.04 | 1098 | 0.11^**^ | 1098 | -0.01 | 1093 | -0.11^**^ | 1078 | 0.02 | 1091 | 0.02 | 1092 | 0.03 | 1090 | 0.02 | 1089 |
| Physical activity: Light | 0.13^**^ | 792 | 0.05 | 792 | -0.07^*^ | 792 | -0.14^**^ | 782 | -0.04 | 792 | 0.02 | 792 | 0.07^*^ | 790 | 0.08^*^ | 788 |
| Physical activity: Light (imputed) | 0.12^**^ | 1098 | 0.07^*^ | 1098 | -0.06^*^ | 1093 | -0.14^**^ | 1078 | -0.03 | 1091 | 0.03 | 1092 | 0.07^*^ | 1090 | 0.09^*^ | 1089 |
| Physical activity: Moderate | -0.17^**^ | 792 | -0.14^**^ | 792 | 0.12^*^ | 792 | 0.24^**^ | 782 | 0.06 | 792 | -0.11^*^ | 792 | -0.16^**^ | 790 | -0.09^*^ | 788 |
| Physical activity: Moderate (imputed) | -0.16^**^ | 1098 | -0.13^**^ | 1098 | 0.16^**^ | 1093 | 0.22^**^ | 1078 | 0.11^**^ | 1091 | -0.08^*^ | 1092 | -0.16^**^ | 1090 | -0.08^*^ | 1089 |

Note: *p<0.05; **p<0.001
